# Epidemiological and clinical characteristics of COVID-19 in Brazil using digital technology

**DOI:** 10.1101/2020.09.30.20204917

**Authors:** Faissal N. Hajar, Miguel M. Fernandes-Silva, Gustavo S. Pereira da Cunha, Ali B. Hamud, Geny H. Herrera, Valderílio F. Azevedo

## Abstract

**Background:** Brazil has the third-highest number of Coronavirus disease 2019 (COVID-19) cases worldwide. Understanding the epidemiology of COVID-19 from reported cases is challenging due to heterogeneous testing rates. We estimated the number of COVID-19 cases in Brazil on a national and regional level using digital technology.

**Methods:** We used a web-based application to perform a population-based survey from March 21^st^ to August 29^th^, 2020 in Brazil. We obtained responses from 243 461 individuals across all federative units, who answered questions on COVID-19-related symptoms, chronic diseases and address of residence. COVID-19 was defined as at least one of the following: fever, cough, dyspnea and nasal flaring, associated with a history of close contact with a suspect or confirmed COVID-19 case in the previous 14 days. A stratified two-stage weighted survey analysis was performed to estimate the population level prevalence of COVID-19 cases.

**Results:** After calibration weighing, we estimated that 10 339 461 cases of COVID-19 occurred, yielding a 2.75 estimated infection per officially reported case. Estimated/reported ratios varied across Brazilian states and were higher in states with lower human development indexes. Areas with lower income levels displayed higher rates of COVID-19 cases (66 vs 38 cases/1000 people in the lowest and highest income strata respectively, p<0.001), but presented lower rates of COVID-19 testing.

**Conclusion:** In this population-based survey using digital technology in Brazil, we estimated that the COVID-19 case rates were 2.75 times higher than officially reported. The estimated per reported case ratios were higher in areas with worse socioeconomic status.

## INTRODUCTION

The severe acute respiratory syndrome coronavirus 2 (SARS-CoV-2) is the causal agent of the Coronavirus disease 2019 (COVID-19) global pandemic. This infectious disease was first reported in Wuhan, China, in December 2019 and rapidly spread to countries all over the world. The World Health Organization (WHO) declared COVID-19 a global pandemic on March 11, 2020, and approximately twenty-five million people have been infected by August 29, 2020.[1] With high transmissibility and absence of an effective antiviral therapy or vaccine, the COVID-19 pandemic has demanded a rapid public health response to implement effective methods of disease tracking to mitigate and contain the disease spread.

Although COVID-19 case reporting requires accurate testing using RT-PCR or serology, widespread testing has shown to be impractical, with highly heterogeneous per-capita testing rates across different regions in the world, depending on test production inputs, resources and allocation constraints.[2] On the other hand, suspected COVID-19 cases can be identified based on patient-reported symptoms and history of exposure to SARS-COV2, which can be useful in a pandemic scenario, particularly in settings with test shortage.[3] This approach can be implemented on a large scale using mobile apps and web-applications, capable of reaching thousands of patients within days. Indeed, digital technology has been used in the COVID-19 pandemic response in different ways, such as screening of infection, contact tracing, predicting clinical outcomes and providing capacity for virtual care.[4] Digital technology initiatives have been successfully applied in the United Kingdom, United States, Germany and other countries, helping to track the disease spread in real time for a fraction of the cost and with reasonable accuracy.[5-7] However, it is unknown whether digital health technologies can help in developing countries, which face several challenges imposed by socioeconomic factors, lack of availability of digital devices and limited access to the internet.

We, therefore, described the results from the largest web-based collaborative COVID-19 application in Brazil - called Together Against COVID. We used the Together Against COVID database to estimate the cumulative incidence rates of COVID-19 cases in each Brazilian federative unit, and compared them with the respective rates of reported cases. We also evaluated the COVID-19 case rates according to neighborhood socioeconomic characteristics. These results will help provide insights on the epidemiology of the COVID-19 pandemic in Brazil, the world’s third most-hit country and the current epicenter of the pandemic.

## METHODS

### Study Design and Population

This was a nationwide, large survey-based study including all brazilian federative units. On March 21st, 2020, we started a non-governmental web-based application called *Juntos Contra o COVID* - Together Against COVID in Portuguese, available in juntoscontraocovid.org. After consenting, each participant filled in a form answering questions on current symptoms, chronic diseases and address of residence. When they completed the form, they had access to a map showing the location of suspected COVID-19 cases. Participants received a weekly email to update their symptom status.

The survey had no pre-specified eligibility criteria, ranging from all ages. No symptoms nor prior known contact with the coronavirus was used as an inclusion criteria. Furthermore, no exclusion criteria were predefined. Participants from all over the country were able to contribute in a collaborative fashion. For this study, we presented data collected up to August 29th, 2020.

### Recruitment Strategy

The primary recruitment method was the snowball strategy - where previous participants invite known potential participants to enter the survey. This organic sampling method was followed by social media engagement and news portals sharing. However, the latter served, primarily, as a focus for the snowball recruitment strategy.

Two main strategies were used to enhance patient recruitment. The first relied on social media and message platforms. Predetermined messages and images were created to publicize the platform as a free, collaborative and useful tool - where people could navigate through a map and check if their surroundings had suspected COVID-19 cases. This strategy worked as a mainstream method to enhance individuals’ access to the web-platform, where they were invited to participate in the research project, sharing their symptoms and health information. The second strategy leaned on publicizing the platform in nationwide news channels. The overall reach of the platform was over 5 million Brazilians.

### Clinical and Socioeconomic Covariates

We collected data on self-reported age, gender, COVID-19 test result, fever, cough, dyspnea, ageusia, anosmia, comorbidities (diabetes, high blood pressure, cardiovascular disease, pulmonary disease, renal disease, rheumatic disease, cancer and chronic corticoid use), close contact with suspected COVID-19 case, close contact with confirmed COVID-19 case, travel history to known COVID-19 community transmission area and residence address. Each participant’s home address was geocoded, and matched to the respective census block from the Brazilian Institute of Geography and Statistics (IBGE) database.[8] We obtained each census block average income and number of individuals per household from the 2010 Brazilian census.[8] Income level was displayed in 2010 Brazilian minimum wage (equivalent to US$ 377.00).

### Suspected COVID-19 case definition

We defined suspect COVID-19 cases based on the Brazilian Health Ministry’s definition, which is similar to the WHO’s suspect case definition for global surveillance.[9, 10] Accordingly, there are three scenarios where a patient is a suspected COVID-19 case:

I. Patient evolves with fever and at least one of the following signs and symptoms: cough, dyspnea, nasal flaring; and travel history to local transmission area, according to WHO, up to 14 days before the first symptoms; or
II. Patient evolves with fever and at least one of the following signs and symptoms: cough, dyspnea, nasal flaring; and close contact with a suspected COVID-19 case, up to 14 days before the first symptoms; or
III. Patient evolves with fever or at least one of the following signs and symptoms: cough, dyspnea, nasal flaring; and close contact with confirmed COVID-19 case, up to 14 days before the first symptoms;

A participant who filled in any of the above rules was defined as a suspected COVID-19 case.

### Data Management

All data were maintained on an AWS hosted Structured Query Language (SQL) database. Participants were identified by an anonymous hash value created upon form completion. This value was used to track multiple responses from the same user given a pre-established email pattern.

### Statistical Analysis

We performed a stratified two-stage weighted survey analysis to estimate the population level prevalence of COVID-19 cases. A Federative unit was used as stratum, the census block as the first stage (primary) sampling unit, and individuals as the second-stage sampling unit. Sampling weights were defined by the inverse of the probability of at least one census block response within each federative unit and the inverse of the probability of the individual response within the respective census block. To account for selection bias from survey responses, sampling weights were adjusted using the general regression method for calibration with age, sex and census block average income level (< ½, ½ to <1, 1 to <2, 2 to <3, 3 to <5, and 5 and above minimum wage) as auxiliary variables. Age- and sex-specific population totals were obtained from the 2020 estimated population size for each federative unit according to the IBGE.[8] Income level category-specific population totals were obtained from the 2010 Census data. The model performance was checked by comparing the final estimated average age, sex and income levels from the survey analysis with official population data from IBGE Census.

From this model, we estimated the cumulative incidence rates of COVID-19 cases by federative unit and compared them with the reported cases. Finally, we performed a logistic regression model using the same survey-based approach to evaluate the association of income level and number of individuals per household (< 2, 2 to < 3, and 3 and above) with the rates of COVID-19 cases. The analyses were performed using Stata version 15.1 (Stata Corp, College Station, TX).

## RESULTS

From March 21 to August 29, we obtained responses from 243 461 individuals in 3277 cities in Brazil, including all the 26 states and the Federal District (additional info in Supplementary table 1). Among participants with suspected COVID-19, the most common symptom was fever (76%) followed by cough (75%), anosmia (40%), ageusia (40%) and dyspnea (38%), and 44% of them reported contact with a confirmed COVID-19 case in the previous 14 days (Supplemental figure 1).

**Table 1.** Demographic characteristics of the sample participants and the Brazilian population and changes in estimates after weighing and calibration. The table shows the demographic characteristics of the sample participants and the Brazilian population as well as changes in estimates after performing weighing and calibration of the sample data.

|  | Sample (Unweighted) | Weighted | Calibrated | Population |
| --- | --- | --- | --- | --- |
|  | **n=243461** |  |  | **N=211755692** |
| Age in years, mean | 42.2 | 40.3 | 36.3 | 34.5 |
| Women, % | 59.6% | 57.1% | 51.1% | 51.1% |
| **Age stratum, men** |  |  |  |  |
| < 20 years | 1.2% | 1.8% | 14.5% | 14.5% |
| 20 to < 40 years | 17.7% | 20.3% | 16.1% | 16.1% |
| 40 to < 60 years | 15.5% | 15.7% | 12.0% | 12.0% |
| 60 to < 80 years | 5.9% | 5.0% | 5.5% | 5.5% |
| 80 years or older | 0.1% | 0.1% | 0.8% | 0.8% |
| **Age stratum, women** |  |  |  |  |
| < 20 years | 2.0% | 2.7% | 13.9% | 13.9% |
| 20 to < 40 years | 26.7% | 27.5% | 16.2% | 16.2% |
| 40 to < 60 years | 22.8% | 20.6% | 13.0% | 13.0% |
| 60 to < 80 years | 8.0% | 6.2% | 6.7% | 6.7% |
| 80 years or older | 0.2% | 0.1% | 1.3% | 1.3% |
| Income per capita in BRL, mean | 1709.46 | 1114.36 | 717.57 | 736.00 |
| **Income stratum (in minimum wage)\*** |  |  |  |  |
| < 1/2 | 1.3% | 6.6% | 29.2% | 20.9% |
| 1/2 to < 1 | 16.3% | 32.2% | 38.1% | 41.1% |
| 1 to < 2 | 35.6% | 36.2% | 23.5% | 25.4% |
| 2 to < 3 | 17.9% | 10.8% | 4.6% | 5.0% |
| 3 to < 5 | 17.0% | 8.7% | 2.9% | 3.2% |
| 5 to < 10 | 11.2% | 4.3% | 1.5% | 1.7% |
| 10 or over | 0.8% | 0.2% | 0.1% | 0.1% |

\*Income was obtained at the census block level from 2010 Census data

**Figure 1.**
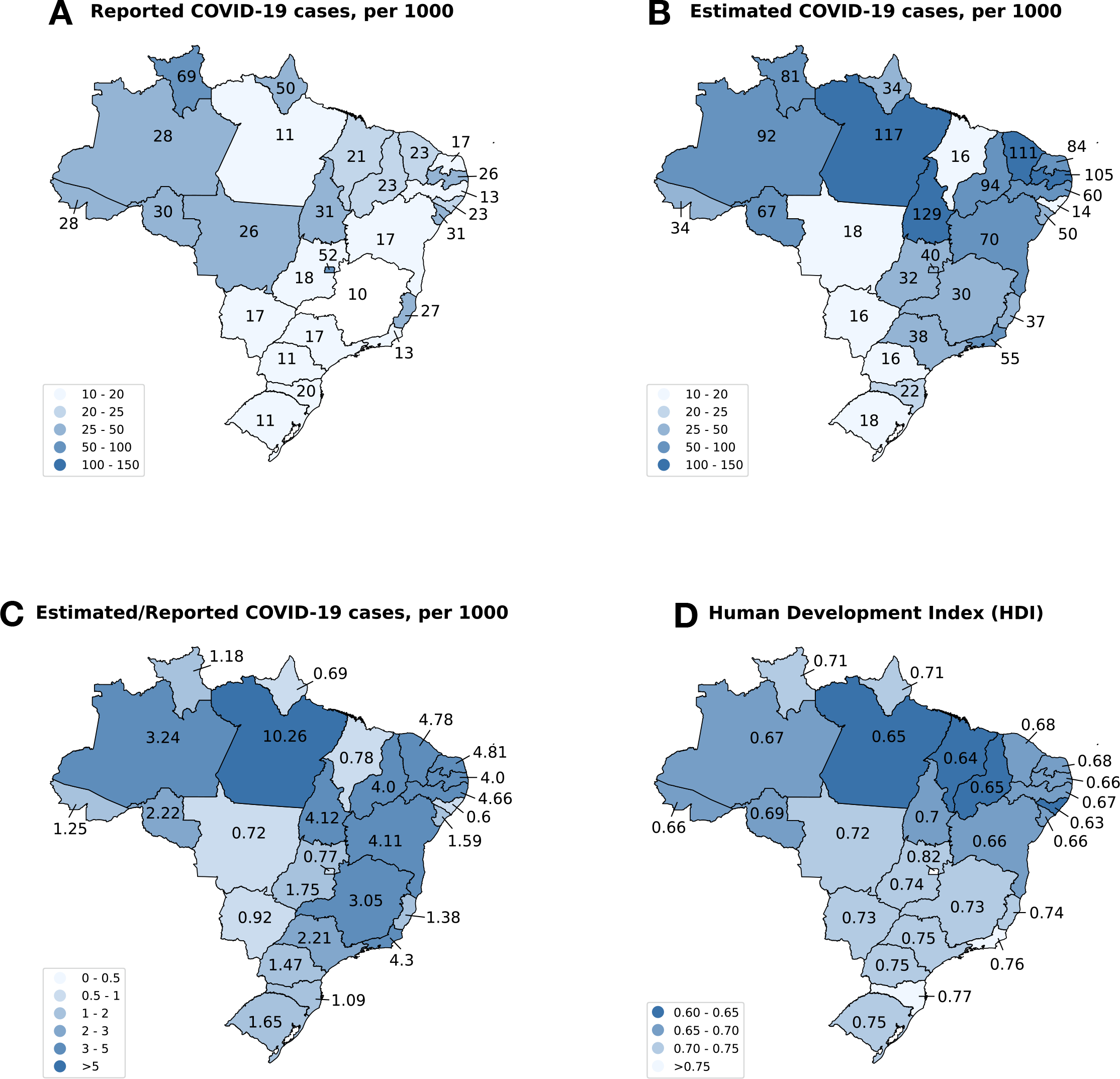
Distribution by federative unit of COVID-19 reported and estimated cases. Distribution of officially reported (A) and survey-based estimated (B) COVID-19 cases per 1000 inhabitants according to the federative unit, ratios between reported and estimated cases (C) and the Human Development Index (D) according to the federative unit. The highest rates of COVID-19 cases were observed in the North and Northeastern regions, but the number of estimated cases was higher than reported in all federative units. The estimated/reported COVID-19 ratios were higher in the Northeastern region, particularly in states with lower Human Development Indexes.

Table 1 shows the descriptive characteristics of the sample, along with data after survey weighting and calibration methods, and the population demographics according to Brazilian census.[11] Compared with Brazilian population, our sample had an over-representation of women between 20 and 60 years old, and an under-representation of individuals younger than 20 years old and older than 80 years old. Moreover, it was over-represented by individuals living in regions with higher income per capita. After weighing and calibration, the estimated parameters from the sample became similar to the population data according to Brazilian census.[11]

By August 29, 2020, the Ministry of Health reported 3,761,391 COVID-19 cases, as displayed in Figure 1A. Nevertheless, we estimated that 10,339,461 cases occurred in the same period, with different relative distributions among the federative units (Figure 1B, table 2). Overall, the ratio between the estimated and reported COVID-19 cases was 2.75:1 in Brazil, but this ratio highly varied across states, with higher ratios in the states of Pará (PA), Rio Grande do Norte (RN) and Pernambuco (PE) (Figure 1C). In Mato Grosso (MT), Mato Grosso do Sul (MS), Maranhão (MA), Amapá (AP) and Alagoas (AL) we estimated fewer cases than officially reported (Figure 1C). The figure 1D displays the Human Development Indexes (HDI) in each state according to the Brazilian Institute of Geography and Statistics (IBGE)[8] with the lowest indexes in the North and Northeast regions. State-level estimated/reported COVID-19 case ratios appeared to be inversely associated with HDI and COVID-19 testing rates, with higher ratios in states with lower HDI and lower testing rates (Figure 2).

**Table 2.**
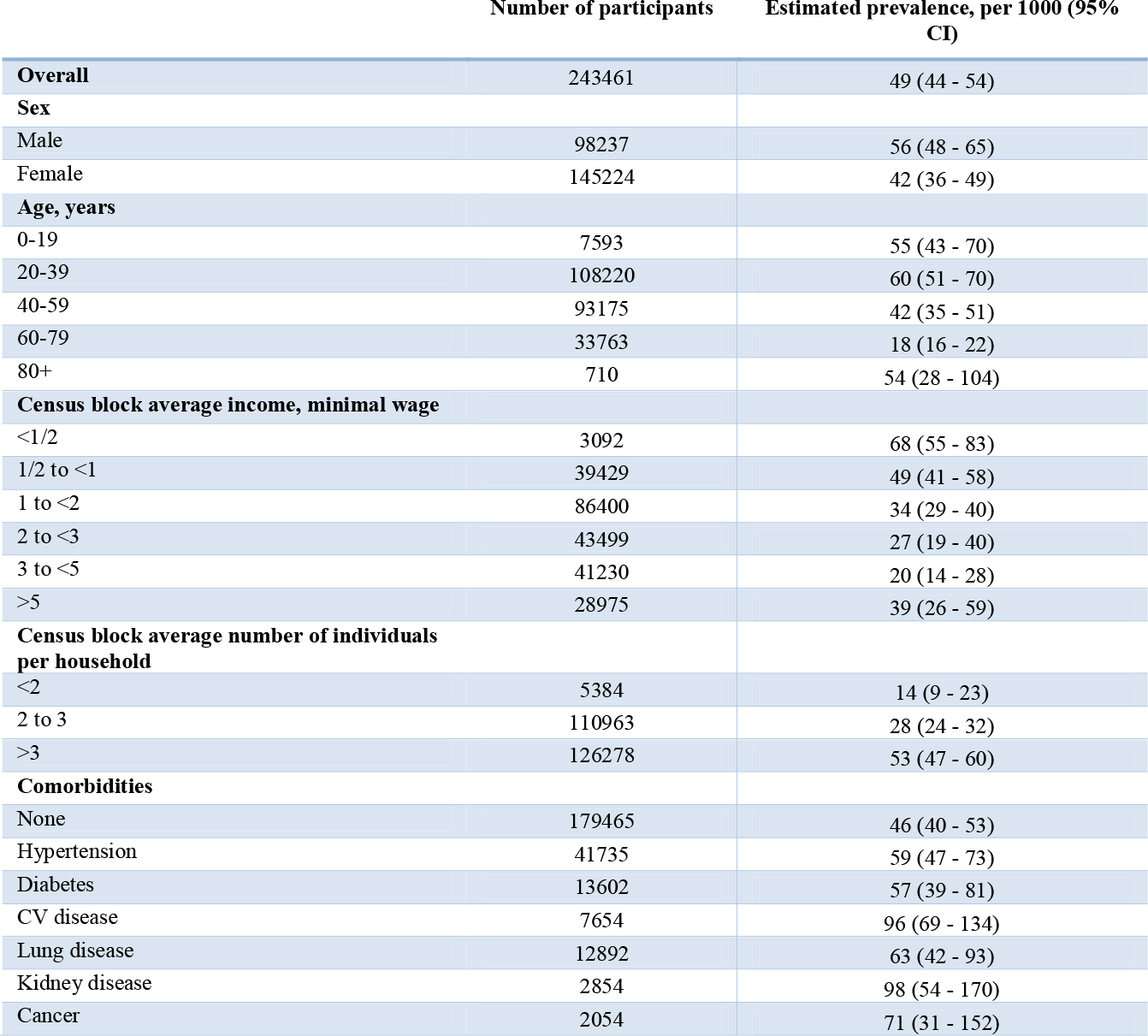
Estimated prevalence of COVID-19 in the Brazilian population according to demographic characteristics and comorbidities. The table shows the estimated prevalence of COVID-19 in the Brazilian population according to sex, age, Census block average income and average number of individuals per household and comorbidities. *CV disease, Cardiovascular disease*.

**Fig. 2.**
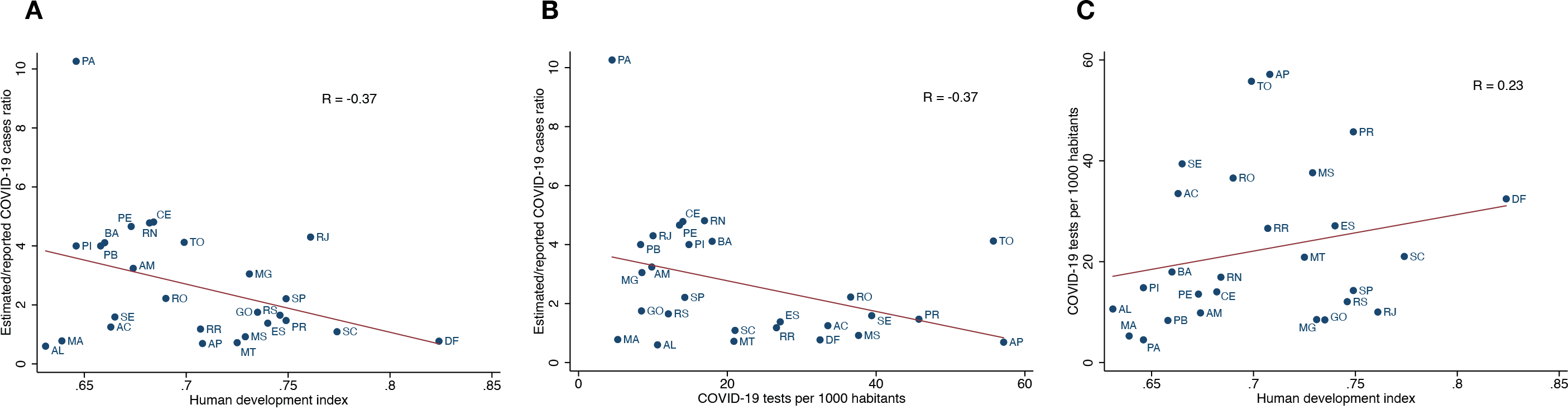
Association per federative unit between estimated/reported COVID-19 cases ratio, COVID-19 tests per 1000 habitants and Human Development Index. The solid line represents the linear regression between the variables. Figure 2A shows the relation between estimated/reported COVID-19 cases ratio and COVID-19 tests. Figure 2B shows the relation between estimated/reported COVID-19 cases ratio and Human Development Index. Figure 2C shows the relation between COVID-19 tests and Human Development Index. *AC, Acre; AL, Alagoas; AM, Amazonas; AP, Amapá; BA, Bahia; CE, Ceará; DF, Distrito Federal; ES, Espírito Santo; GO, Goiás; MA, Maranhão; MG, Minas Gerais; MS, Mato Grosso do Sul; MT, Mato Grosso; PA, Pará; PB, Paraíba; PE, Pernambuco; PI, Piauí; PR, Paraná; RJ, Rio de Janeiro; RN, Rio Grande do Norte; RO, Rondônia; RR, Roraima; RS, Rio Grande do Sul; SC, Santa Catarina; SE, Sergipe; SP, São Paulo; TO, Tocantins*.

The COVID-19 case rates in Brazil were significantly associated with socioeconomic factors, showing a “J” shaped association with income, with a down trend from the lower to the upper-middle income regions and an uptrend from upper-middle (3 to < 5 minimum wage) to the upper income stratum (5 and over minimum wage, figure 3A). The estimated COVID-19 rates in the lower income stratum were approximately three times higher than the rates in the upper-middle income stratum (68 vs 20 per 1000 people) and twice the one in the upper income stratum (68 vs 39 per 1000 people, Figure 3A). On the other hand, self-reported COVID-19 testing was the most common in the highest income stratum (123 tests per 1000 people) compared to other strata (p<0.001, Figure 3B).

**Fig. 3.**
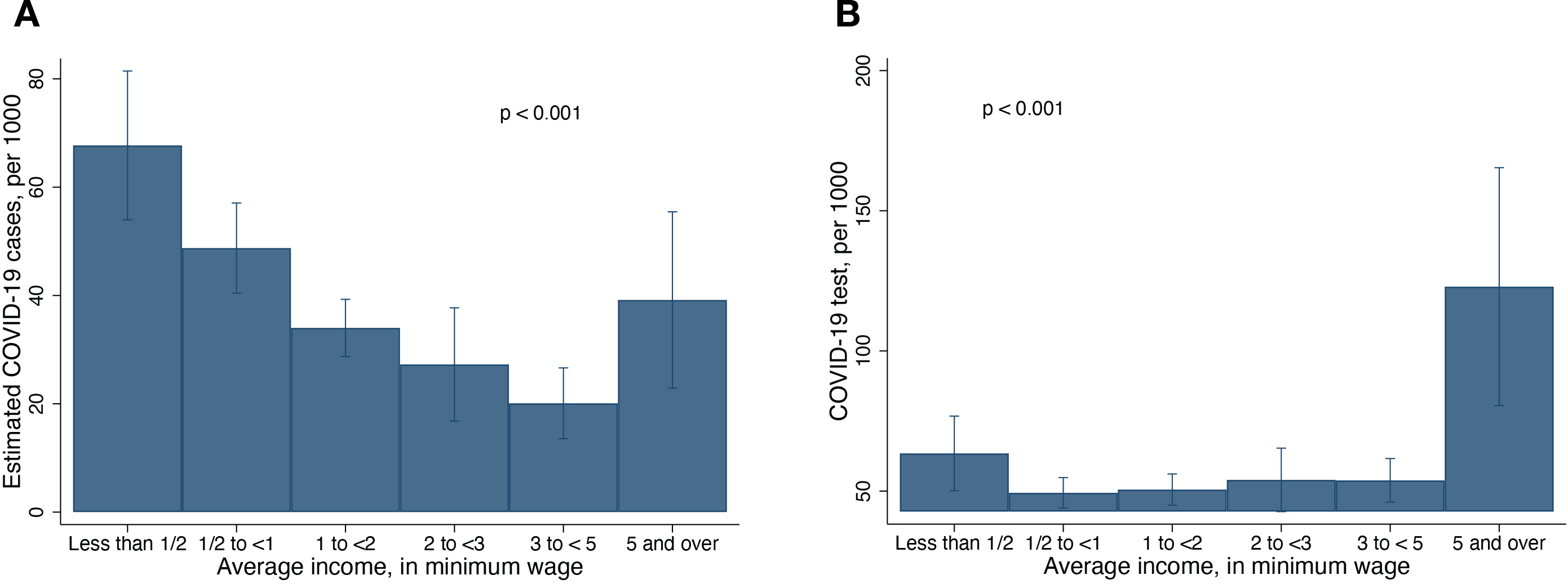
Estimated COVID-19 cases and tests by income strata. Estimated cumulative incidence rates of COVID-19 cases (A) and self-reported COVID-19 testing (B) according to census block income level. While the COVID-19 rates were inversely associated with income level, self-reported COVID-19 testing was considerably higher in the highest income stratum, as compared with all other income strata.

While the COVID-19 case rates were inversely associated with census-block average income [Odds Ratio (OR) 0.74, 95% Confidence Interval (CI): 0.67, 0.82, p<0.001) per category increase], they were directly associated with the average number of individuals living in the same household (OR 1.94, 95% CI: 1.61, 2.36, p<0.001, per category increase, Table 2). In a multivariate analysis, the association between COVID-19 case rates and income remained similar after adjusting for number of individuals per household (OR 0.78, 95% CI 0.69, 0.87, p<0.001)

The COVID-19 rates varied across age strata, with the highest rates in the population between 20 and 39 years and the lowest ones between 60 and 79 years old. Noteworthy, COVID-19 was more common among individuals with comorbidities, particularly those with self-reported cardiovascular diseases, than those with no comorbidities (Table 2).

## DISCUSSION

To our knowledge, this is the first study to estimate the cumulative incidence rates of COVID-19 using a web-based collaborative tool in Brazil. In this population-based survey, the estimated COVID-19 case rate was 2.75 times higher than it has been reported. Under-reporting of COVID-19 varied across states, as reflected by different estimated/reported COVID-19 ratios, and it appears to be higher in states with lower HDI. The rates of COVID-19 cases were higher in neighborhoods with lower income levels, but the rates of self-reported COVID-19 testing was almost twice higher in the highest income compared with the lowest income neighborhoods. Our study shows how digital technology can help understand the epidemiology of COVID-19 in Brazil, which could be replicated in other developing countries. Better understanding of the factors leading to higher case rates among lower income communities can help inform public authorities to better allocate resources and develop strategies to mitigate the spread of COVID-19.

Previous studies used digital technology to track COVID-19 cases based on self-reported symptoms. A multinational Consortium involving a digital health care company and a multidisciplinary team of scientists in the United States and the United Kingdom developed a mobile app to provide real-time epidemiology of COVID-19 during the outbreak.[12] Data from this app have shown that a symptom-based prediction model could estimate the time changes in the incidence of COVID-19 cases days before the officially reported cases.[12] Other countries in Asia and Europe have also implemented mobile apps for different purposes, such as identification of potential cases and tracing contacts of COVID-19 patients to recommend self-isolation.[13] Our study adds that digital technology can be combined with official data to detect the spread of COVID-19 earlier and identify the most affected areas.

Previous studies also suggested under-reporting of COVID-19 cases using sero-prevalence and severe acute respiratory infection (SARI) data.[14-17] A population-based study in Southern Brazil estimated that, for each reported case, there were approximately 10 infections, as reflected by the prevalence of individuals who tested positive for anti-SARS-CoV-2 IgG antibodies, similar to what has been found in other countries.[15,17] Asymptomatic cases - 15% to 40% of COVID-19 infections - can only partially account for these differences.[14] A study from the Sistema de Informação de Vigilância Epidemiológica da Gripe (SIVEP-Gripe) system found an increase in SARI cases with unknown etiology in Brazil from March to May, 2020 that were 2.3-fold higher than the SARI due to COVID-19.[18] In the Metropolitan Region of São Paulo in the southern of Brazil, confirmed COVID-19 diagnosis among SARI cases was less likely in areas with lower per-capita income. Similarly, our study found that COVID-19 case rates from self-reported symptoms were 2.75-fold higher than the reported cases, with higher estimated/reported ratios in states with lower HDI and lower testing rates.[18] This may imply non-equitable access to testing and diagnosis, but the reasons for under-reporting of COVID-19 are complex and multifactorial, involving both healthcare delivery, cultural aspects and socioeconomic characteristics.

We showed a significant association between COVID-19 case rates and socioeconomic factors. Areas with lower income and more people living in the same house were associated with higher rates of COVID-19. Social distancing may be seemingly impossible in small, overcrowded and poorly ventilated houses with a single room, enhancing the transmission of respiratory viruses whenever a family member develops symptoms.[19-21] Although there is a lack of data on the transmission of SARS-Cov-2 in poor communities, a study suggested that a hypothetical pandemic of a new strain of influenza would have greater impact in a low-income country, such as Papua New Guinea, than in a developed country, mainly due to a larger number of individuals per household.[19] Consistent with this hypothesis, the prevalence of COVID-19 was not associated with income strata and number of individuals per household in the seroprevalence study in Spain.[14] This suggests that the higher transmissibility in crowded households may particularly affect lower-income countries. Policy strategies that address crowded households when an individual is tested positive can be important tactics to mitigate the spread of COVID-19 in low-income settings. On the other hand, our data showed that the association between COVID-19 rates and income remained almost unchanged after adjusting for the number of individuals per household, suggesting that other factors play an important role in this association.

We found that the COVID-19 case rates were higher, but testing rates were lower, among census blocks with lower average income. This underscores the heterogeneous access to healthcare even when there is universal health coverage. Individuals in the lower income strata, who were at higher risk of COVID-19, were less likely to receive testing and, therefore, to self-isolate.[22] In addition to the disease itself, poor communities face difficulties indirectly caused by the pandemic, such as a drastic reduction in income and a loss of community targeted support.[23] Besides that, low-income people may be more reluctant to social isolation due to: family responsibilities as a food and wage provider; less flexible jobs; fear of losing their position and lack of formal jobs.[24-26]

Digital technology can be a powerful tool to quickly provide valuable epidemiological information at a low cost. Disease tracking, contact tracing, diagnosis support, clinical status monitoring and telemedicine are among applications that have been implemented in different countries to fight against this pandemic.[4] The impact of the COVID-19 pandemic will be worse in low income settings with weaker healthcare systems, which will increase the world health disparities due to the effect of the negative social determinants. Despite the concerns on limited access to the internet, we demonstrated that low-cost digital initiatives coupled with effective social engagement can be extremely useful to provide important data on disease activity in a developing country with continental proportions.

Our study has limitations that deserve attention. First, we used a web-based application that relies on spontaneous survey responses. Although access to the internet has been reported in 79% of households in Brazil, our sample was under-represented by elderly and low-income individuals.[30] Appropriate weighing and calibration helped mitigate this limitation, as estimated demographics became similar to census-based population data. Second, most COVID-19 cases were defined by self-reported symptoms and history of contact with a suspected/confirmed case, which may overestimate the number of symptomatic infections and does not account for asymptomatic infections. Nevertheless, this strategy is a feasible approach to evaluate large population-based surveys, particularly in settings with insufficient tests for screening every suspected case, such as most regions in the world during this pandemic. Finally, a noteworthy aspect of this initiative was its collaborative and non-profit nature. Over 30 professionals and over 10 companies from various fields gathered resources and expertise to help brazilians stay aware of their surroundings while also contributing to science. This study proves that it is possible to conduct nationwide high-level research in developing countries with limited resources.

### What is already known in this subject?

- Developed countries have successfully used digital technologies to gain insights about epidemiological and clinical data regarding the COVID-19 pandemic
- There is limited evidence on the use of such technologies in developing countries.

### What this study adds?

- More than 240.000 patients were assessed for COVID-19 symptoms and epidemiological data in Brazil using a low cost digital web-application.
- It proved to be a safe and viable method in developing countries, such as Brazil, enabling insights on under notification and the impacts of wage on infection and testing.

## Data Availability

The data underlying this article are available in the article and in its online supplementary material.

## ETHICS

The study protocol was approved by the Ethics Committee from the Federal University of Paraná (CEP CHC/UFPR #35028620.0.0000.0096) as a safe design, offering no harm to any participant. All participants provided consent for non-commercial use of their data.

## ACKNOWLEDGMENTS

We would like to thank Unimed Curitiba for the donation to support the web-based application.

