## Supplemental material for "Epidemiological and clinical characteristics of COVID-19 in Brazil using digital technology"

**Supplementary Material**

**Figure S1. Proportion of self-reported symptoms and positive COVID-19 test among suspected COVID-19 cases.**

**Figure S1.** The figure shows the proportion of self-reported symptoms and positive COVID-19 test among participants defined as a suspected COVID-19 case. The most common symptom was fever (76%) and the least common one was dyspnea (44%). A total of 44% of the participants labeled as suspected COVID-19 case based on the World Health Organization's criteria tested positive to COVID-19.

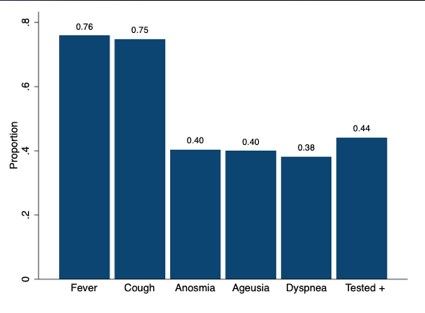

**Table S1. Sample and population number of individuals and census block according to federative unit**

| State | Population | Sample | Sample/population (per 100,000) | Population census block | Sample census block | Sample/Population census block ratio |
| --- | --- | --- | --- | --- | --- | --- |
| AC | 894470 | 159 | 0,018% | 874 | 82 | 9% |
| AL | 3351092 | 683 | 0,020% | 3724 | 381 | 10% |
| AM | 4207714 | 521 | 0,012% | 5644 | 309 | 5% |
| AP | 861773 | 161 | 0,019% | 810 | 91 | 11% |
| BA | 14930424 | 4550 | 0,030% | 23795 | 1887 | 8% |
| CE | 9187886 | 3342 | 0,036% | 13278 | 1798 | 14% |
| DF | 3052546 | 7247 | 0,237% | 4405 | 1689 | 38% |
| ES | 4064052 | 2476 | 0,061% | 6377 | 1176 | 18% |
| GO | 7116143 | 2948 | 0,041% | 9413 | 1402 | 15% |
| MA | 7114598 | 413 | 0,006% | 8803 | 309 | 4% |
| MG | 21292666 | 27439 | 0,129% | 32568 | 9786 | 30% |
| MS | 2809394 | 1709 | 0,061% | 4191 | 772 | 18% |
| MT | 3526220 | 1051 | 0,030% | 5944 | 600 | 10% |
| PA | 8690745 | 1005 | 0,012% | 8776 | 643 | 7% |
| PB | 4039277 | 990 | 0,025% | 5549 | 570 | 10% |
| PE | 9617072 | 2029 | 0,021% | 12379 | 1124 | 9% |
| PI | 3280697 | 494 | 0,015% | 5251 | 237 | 5% |
| PR | 11516840 | 29536 | 0,256% | 17475 | 6442 | 37% |
| RJ | 17366189 | 23921 | 0,138% | 27774 | 8691 | 31% |
| RN | 3534165 | 1733 | 0,049% | 4290 | 775 | 18% |
| RO | 1796460 | 437 | 0,024% | 2345 | 236 | 10% |
| RR | 631181 | 99 | 0,016% | 824 | 71 | 9% |
| RS | 11422973 | 15680 | 0,137% | 22342 | 5344 | 24% |
| SC | 7252502 | 17449 | 0,241% | 11885 | 4384 | 37% |
| SE | 2319032 | 704 | 0,030% | 3298 | 365 | 11% |
| SP | 46289333 | 96462 | 0,208% | 66213 | 27144 | 41% |
| TO | 1590248 | 223 | 0,014% | 2101 | 135 | 6% |

**Table S1**. The table presents the sample size and population number of individuals for each federative unit, as well as the census block equivalents. Thus, the most represented federative unit was SP (41%) and least represented one was AM (5%). This analysis was performed before weighing and calibration. *AC, Acre; AL, Alagoas; AM, Amazonas; AP, Amapá; BA, Bahia; CE, Ceará; DF, Distrito Federal; ES, Espírito Santo; GO, Goiás; MA, Maranhão; MG, Minas Gerais; MS, Mato Grosso do Sul; MT, Mato Grosso; PA, Pará; PB, Paraíba; PE, Pernambuco; PI, Piauí; PR, Paraná; RJ, Rio de Janeiro; RN, Rio Grande do Norte; RO, Rondônia; RR, Roraima; RS, Rio Grande do Sul; SC, Santa Catarina; SE, Sergipe; SP, São Paulo; TO, Tocantins.*
